# Brain Impairment revealed by Multi-Modality MRI in Parkinson’s Disease

**DOI:** 10.1101/2020.06.22.20136663

**Authors:** Zhang Ran, Gong Ping, Ge Haitao

## Abstract

**Objective:** To study the abnormal brain regions of patients with Parkinson’s disease (PD) using multimodality MRI to provide complementary information for early detection for PD.

**Methods:** 27 patients with early PD and 25 normal ageing volunteers were included in the study. Multimodality MRI data were acquired and processed to extract neuroimaging features to test the structural and functional changes using a two-sample t-test.

**Results:** The changes of brain regions were disagreed for different modality MRI data between PD and normal ageing individuals. Nevertheless,the postcentral gyrus, precentral gyrus, lingual gyrus and paracentral lobule were significantly different for all three modalities.

**Conclusion:** Multimodality MRI data can reflect the structural and functional changes of PD, and reveal the hidden information which is of great significance to assist early detection for PD.

## Introduction

Parkinson’s disease (PD) is a common neurodegenerative disease, of which the average age of onset is around 60-year-old. The primary pathological changes of PD are the degeneration and death of dopaminergic neurons in the substantia nigra and the formation of Lewy bodies. PD is characterized by four primary motor symptoms, such as resting tremor, rigidity, bradykinesia and postural instability. It is challenging to use the routine imaging examination for the early-detection of PD due to the pathological changes often occurring prior to the brain impairment. Therefore, motor symptoms are usually adopted as diagnostic indicators for PD in clinical application. Consequently, it is generally developed to the middle or late stage when PD was confirmed. Thus, majority of PD patients missed the best opportunity for early diagnosis and treatment. It will be greatly beneficial for PD patients if it is early-diagnosed at early stage and treated with effective neuroprotective therapy. Therefore, it is of great significance to seek effective imaging biomarkers to assist the early diagnosis of PD.

MRI has been greatly developed in recent decades and turns out to be extremely helpful for the early diagnosis of PD. MRI is used to extract neuroimaging features, which will help to assist early diagnosis of PD and provide a new perspective for early detection. T1-weighted images will provide the anatomical information of brain, particularly for the subcortical nucleus. It has been reported in the literature that PD patients exhibited morphological changes for the density, volume of gray/white matter and cortical thickness using voxel-based morphometry (VBM) approach. Diffusion tensor imaging (DTI) takes advantage of the difference of diffusion movement of water molecules to directly reflect the direction of fiber in the white matter of the brain. DTI-derived measures can be used to evaluate the fiber integrity of the brain for PD, such as fractional anisotropy (FA), mean diffusivity (MD), radial diffusivity (RD) and axial diffusivity (AD). Functional magnetic resonance imaging (fMRI) indirectly measures the fluctuation of blood oxygen level-dependent (BOLD) signals, which reflects the spontaneous neural activity of the brain in the resting state. Certain metrices have been widely used in both healthy subjects and patients with brain disorders, such as regional homogeneity (ReHo), amplitude of low-frequency fluctuation (ALFF), fractional amplitude of low-frequency fluctuation (fALFF), and seed-based correlation analysis (SCA), independent component analysis (ICA) and connectome analysis based on graph theory. Numerous neuroimaging features have been shown to have the potential to characterize and classify various brain disorders with a single-modality MRI. Yet, each MRI modality also has its own unique technological shortcomings. Multi-modality MRI can reveal the unique structural and functional information of the brain from multiple perspective.

In this study, we try to extract neuroimaging features to discriminate PD from normal ageing individuals using multi-modality MRI, including T1, DTI and rest fMRI images. It will provide complementary information for early detection for PD and assist clinical diagnosis and treatment.

## Materials and Methods

### Subjects

A total of 27 PD patients (including 16 males and 11 females with a mean age of 63.2 ±7.82 years) from the Affiliated Hospital of Xuzhou Medical University were recruited from 2015 to 2017. All patients satisfied the UKPDS Brain Bank diagnostic criteria and were assessed by neurologists experienced in diagnosing PD. All patients have/had been reported to be positively respond to levodopa. 25 age-(P=0.72) and sex-matched (P=0.67) normal aged volunteers (13 males and 12 females with a mean age of 60.1±6.45 years) were also included in this study. There are no MRI contraindications, no distinct head trauma, no brain parenchyma lesions, no drug abuse, no alcoholism for all subjects. Permission for this study was obtained from the Xuzhou Medical University research ethical committee and all subjects consented to participation.

### Image Acquisition

Multimodality MRI data were acquired imaging on a 3 T MR scanner (Discovery 750w, GE Healthcare, Waukesha, WI). DTI data were acquired with a single-shot EPI pulse sequence in the axial plane, with repetition time (TR)10s, echo time (TE) 74.2ms; in-plane matrix 128*128; FOV 25.6*25.6 cm, and 3 mm slice thickness. There were 60 diffusion directions with weighting (b) = 1000 s/mm2 and 4 non-diffusion weighted T2 (b0) images. Resting-state fMRI (rsfMRI) data were acquired using a gradient EPI sequence with the following parameters: TR = 2000 ms, TE = 30 ms, 90° flip angle, 22 cm field of view, 64 × 64 in-plane matrix, slice thickness 3.0 mm without gap. Subjects were instructed to keep their eyes open during scanning. To facilitate brain segmentation and co-registration between imaging modalities, a 3D T1-weighted image (sagittal orientation; frequency encoding in the superior-inferior direction; TR/TE = 8.5/3.25 ms; flip angle=15°; FOV=25.6 cm; 256*256 acquisition matrix; and 1.0-mm slices) was also acquired.

### Image Processing

Structural image preprocessing was performed using CAT12 toolbox (http://dbm.neuro.uni-jena.de/cat/) within SPM12 (http://www.fil.ion.ucl.ac.uk/spm). T1-weighted images were corrected for bias-field inhomogeneity using the N4 algorithm. After correction, all images were segmented into gray matter, white matter and cerebrospinal fluid density maps. In addition to segmenting into three different components, T1-weighted images were also used to perform AAL parcellation (78 cortical regions of interest) to define the network nodes

DTI data were preprocessed using PANDA (https://www.nitrc.org/projects/panda/). The preprocessing approaches included skull-stripping, and corrections for head motion and eddy current distortions. Fractional anisotropy (FA) was calculated for each subject after correction.

Preprocessing of rsfMRI data was implemented using DPARSF (http://www.restfmri.net). The first 10 functional volumes were discarded to reach steady state magnetization. The remaining images were slice-timing corrected and realigned for head-motion correction. Head motion was assessed for each subject, and subjects were excluded if the translation of head motion exceeded 3 mm or the rotation exceeded 3 degrees. Subsequently, the images were spatially normalized to the Montreal Neurological Institute (MNI) EPI template, resliced to 3 mm isotropic voxels, and then smoothed with a Gaussian kernel with a full-width at half-maximum (FWHM) of 6 mm. Detrending and band-pass filtering (0.01Hz-0.08 Hz) were then performed to remove high-frequency physiological noise and low-frequency fluctuations. Finally, nuisance covariates including Friston 24 head motion parameters, white matter/cerebrospinal fluid signal were regressed out, and then residual images were saved to compute functional connectivity (FC). The functional connectivity network for each individual was computed by computing the mean time course in each AAL region followed by computation of the Pearson correlation matrix. The resulting correlation matrix was then converted to z-scores by the Fisher transformation.

### Statistical Analysis

Two-sample t-test was used to test the structural and functional changes between PD and normal controls with age and sex as covariates. The correction for multiple comparison was performed using false discovery rate (FDR) with q-value setting to 0.05.

## Results

Voxel-wise analysis of structural images showed the increased density of grey matter in the bilateral hemispheres in PD compared with that in normal ageing subjects, including bilateral orbital superior frontal gyrus, left peri-calcarine fissure cortex, left orbital middle frontal gyrus, right middle temporal gyrus, left middle occipital gyrus, left thalamus, left middle frontal gyrus, left precuneus, left angular gyrus, bilateral precentral gyrus, bilateral postcentral gyrus, left dorsolateral superior frontal gyrus and left paracentral lobule. Decreased density of grey matter of bilateral hemispheres was also found in PD, including bilateral middle temporal gyrus, right lingual gyrus, left superior orbital frontal gyrus, right middle frontal gyrus, left middle occipital gyrus, right medial and parietal cingulate gyrus, left inferior parietal angular gyrus and left postcentral gyrus.

DTI analysis showed that FA values in bilateral hemispheres in PD were increased, including amygdala, hippocampus, rectus gyrus, thalamus, middle occipital gyrus, peri-calcarine fissure cortex, precuneus, medial superior frontal gyrus, medial and paracentric gyrus, middle frontal gyrus, postcentral gyrus and paracentric lobules. FA values in bilateral hemispheres of PD also showed decreased in fusiform gyrus, lenticular pallidus, lenticular pallidus, lingual gyrus, superior temporal gyrus, inferior frontal gyrus, middle occipital gyrus, postcentral gyrus, precentral gyrus, middle frontal gyrus and dorsolateral superior frontal gyrus.

Functional connectivity analysis demonstrated that there are differences in clustering efficiency in precentral gyrus, lingual gyrus, suboccipital gyrus, fusiform gyrus, postcentral gyrus, paracentral lobule, superior temporal gyrus, middle temporal gyrus and inferior temporal gyrus between PD and normal ageing.

## Discussion

Parkinson’s disease (PD) is a progressive neurodegenerative disorder that affects predominately dopaminergic neurons in the pars compacta of substantia nigra, which leads to the decrease of dopamine afferent in the striatum and the abnormality of the cortical-striatal-cortical circuit. Literatures have shown extensive loss of neurons and degeneration of α-synuclein in brainstem and cerebral cortex in PD. The accumulation of α-synuclein hinders the release of neurotransmitters and leads to neuronal necrosis. Thus, PD patients not only suffer from dyskinesia, but emotional and cognitive impairments such as pain, anxiety and depression. Although a great progress has been made in comprehensive understanding of the pathology, pathophysiology, clinical manifestations and diagnostic techniques for PD, it is still lack of robust biomarkers for the early detection of PD. It will provide significant help for the early diagnosis of PD if specific markers based on brain MRI images were found to distinguish PD from normal ageing subjects.

With the rapid development of structural MRI, T1-weighted images are sensitive to the morphological changes of brain structures. It has been reported gray matter volume changes at the early stages of PD using voxel-based morphology approach. Gao et al. found widespread cortical and subcortical atrophy in the frontal lobe, medial temporal lobe, basal ganglia and limbic system in PD. Li et al. showed that the amygdala was the earliest affected cortical region in PD, and the density decreased in bilateral amygdala, right superior temporal gyrus, right fusiform gyrus and parahippocampal gyrus. It also reported that the density of gray matter in the right amygdala was negatively correlated with autonomic nervous dysfunction and positively correlated with cognitive ability. In this study, we also reported that in addition to the brain areas of cortical atrophy, which was in line with the previous studies, there was also some brain areas demonstrated increased gray matter volume. It may be due to the necrosis of massive neurons, which result in impaired information transmission in the limbic system, and compensatory reinforcement of gray matter in other brain regions.

DTI can be used to evaluate the integrity of brain fiber tracts of PD. A large amount of studies has confirmed that the change of FA values of fiber tracts can be used as an imaging marker to predict PD. Lorio et al. found that the FA values of midbrain in PD increased, which was related to the sleep disturbance. Haghshomar et al. performed a systematic review of the published literature and found that PD patients would exhibit up cognitive impairment, facial recognition impairment, olfactory impairment and tremor after the decrease of FA in the lower longitudinal tract and the destruction of the microstructure of the lower longitudinal tract. Zhang et al. reported that the FA values of corpus callosum, bilateral radial corona and left cingulate gyrus in PD with apathy symptoms were significantly decreased than those without apathy symptoms. Besides, the FA values were negatively correlated with the clinical scores with apathy symptoms. In this study, the results showed that the white matter lesions were coincided with gray matter lesions for PD. This is because when PD causes changes in the gray matter of the brain, the white matter in the adjacent lesions will also change accordingly.

Functional MRI is valuable for the hemodynamics changes of brain caused by neural activity, and can be used to capture the information of brain function. When the brain is engaged in information processing and behavioral execution, it can be regarded as a complex dynamic network. Wu et al. found that the clustering efficiency and path length of PD were higher than that of normal ageing, which meant the degree of network collectivization increased and implied part of the brain is attacked. Also, the information transmission for PD became slower. These results indicated that the stability of the small-world network of PD was damaged and the efficiency of information processing was reduced. Sang et al. found the decreased global efficiency of PD, which suggested the functional network of PD tended to be random and disrupted, and the communication of information is limited. Fang et al. concluded that the global and local efficiency of brain network in patients with PD increased, while path length decreased. This indicated that the patients had brain network destruction at the early stage of PD. In this study, we reported that there were clustering efficiency changes in some brain regions of PD. It meant the change in the ability to communicate directly within the brain region for PD.

In this study, we studied the brain structural and functional changes for PD using multi-modality MRI. The results showed the inconsistency between structural and functional changes, which may be related to metabolic changes earlier than morphological changes in the development of PD. Therefore, it is of great significance to seek neuroimaging biomarkers to provide valuable tools for the early diagnosis for PD.

## Data Availability

The data that support the findings of this study are available from the corresponding author upon reasonable request.

## Acknowledges

This study was supported by University Science Research Project of Jiangsu Province (16KJD320006) and Science and Technology Plan Projects of Xuzhou (KC16SL114).

